# Indian community’s Knowledge, Attitude & Practice towards COVID-19

**DOI:** 10.1101/2020.05.05.20092122

**Authors:** Balvir Singh Tomar, Pratima Singh, Supriya Suman, Preeti Raj, Deepak Nathiya, Sandeep Tripathi, Dushyant Singh Chauhan

## Abstract

As COVID-19 pandemic has caused unprecedented human health consequences. Knowledge, attitude, perception of general population of India towards the transmission and prevention plays vital role for effective control measures. The study was conducted to assess the knowledge, attitude and practice of the general public of India on COVID-19. In this study, a web-based cross-sectional survey was conducted between 10^th^ March to 18^th^ April 2020. A 19-item questionnaire was generated, Cronbach’s alpha was used to measure the internal consistency of the questionnaire & randomly distributed among the public using Google forms through social media networks. The chi-square test or Fischer exact test was used to compare categorical data and multiple linear regression was used to identify factor influencing KAP. Among 7978 participants, the overall knowledge, attitude and practice score was 80.64%, 97.33% and 93.8% consecutively. Majority of Indian population demonstrated preceded good knowledge, positive attitude and good practice regarding COVID-19 pandemic.

## INTRODUCTION

The novel coronavirus disease 2019 (COVID-19) caused by Severe acute respiratory syndrome coronavirus 2 (SARS-CoV-2) was Initially diagnosed from Wuhan, Hubei Province (Mainland China), has already taken on pandemic proportions, affecting whole world in the minuscule of time [1,2,3]. As of April 30, 2020, >3.3 million cases of COVID-19 cases had been confirmed resulting in 234,139 deaths worldwide [4]. In reciprocation to the outbreak, World Health Organization (WHO) declared it as a public health emergency of international concern and called global imperative efforts to prevent the escalation [5]. SARS-CoV-2 is an enveloped RNAβ -coronavirus with an outer fringe of envelope proteins resembling like crown, which has a phylogenetic genome similarity with highly pathogenic and transmissible with another known coronavirus i.e. SARS-CoV-1 (2003) and MERS-CoV (2012) [6,7,8,9]. Studies suggest the basic reproduction number (R_0_) of SARS-CoV-2 be around 2.2 or more upto 6 [10], making the virus propagate at an alarming rate and proving to be very expeditious and erratic [11,12].

India is a country of vast socio-cultural diversity; health inequalities and economic disparity presents with challenges and threat by the growing pandemic of COVID-19. Enforcement of immediate lockdown, which was praised by WHO as “tough and timely” and cluster containment to break the chain transmission are effective approach [13,14]. India is the second-largest internet user in the world with >560 million from 1.39 billion gross population [15]. One threat to the COVID-19 response in India is the ubiquitous spread of misinformation by raising falsehoods like rinsing the nose with saline, spraying of alcohol and chlorine or 5G mobile networks inhibiting the spread of the virus, during the crisis is dangerous because it can mislead and confuse the public [16]. Over 3 billion posts and 100 billion interactions are present on COVID-19 making infodemic spread faster than a pandemic [17]. The most important factor in preventing the spread of the virus locally is the empowered citizens with the right information and taking advisories being issued by the Ministry of Health & Family Welfare, Government of India regularly.

To guarantee the final success, people’s adherence to these control measures is essential, which is largely affected by their knowledge, attitudes, and practices (KAP) towards COVID-19 in accordance with KAP theory [18,19].

To address the gap, the study was carried out with the following objectives:

- Primary Objective – To assess the KAP regarding COVID-19 among the general population of India.
- Secondary Objective – To assess the factors associated with the level of KAP regarding the COVID-19 outbreak.

## MATERIALS AND METHODS

### Study design & data collection

This is a cross-sectional survey conducted via snowball sampling technique from 10^th^ March to 18^th^ April 2020 in India. As Community-based survey was not feasible due to nationwide lockdown in India. A 19-item questionnaire was developed using google forms. Information published in literature including publications available on WHO and the Centers for Disease Control and Prevention (CDC) [20,21,22,23] was utilised at large. A Pilot study was conducted to understand the barriers faced by participants and the chronology of the questionnaire. The questionnaire was made available to the participants through emails and social networking platforms such as WhatsApp, LinkedIn, Instagram, and Facebook. The cover page of the questionnaire includes a consent form, a declaration of confidentiality, and anonymity. Participants with age more than 18 years who can understand the content of the survey and willing to participate were included.

### Questionnaire and scoring

The questionnaire consists of two parts-Demographic details and KAP study. Demographic Variables includes; Sex (Male or Female), Age (18-30, 30-50 or > 50 years), Marital Status (Single or Married), Education level (< Senior Secondary, Senior Secondary, Graduate,≥ Post Graduate), Occupation (Unskilled, Skilled, student and unemployed, self-employed, professional), Geographic location (different states of India), Place of residence (urban or rural).

The knowledge section consisted of two parts – 10 questions regarding clinical symptoms, prevention, and control of disease (K1-K10) and 3 questions regarding myth busters on COVID-19 (K11-K13). Each question has three options (Yes/No/Don’t Know). A correct answer was given 1 point and an incorrect answer was given 0 point. Overall knowledge scores ranged from 0-13. Individuals scoring 11 or above were categorized as excellent whereas below 11 scores were brought under poor knowledge.

Evaluation of attitude of the general public was done by 3 Questions (A1-A3) comprising questions assessing viewpoint on social distancing, control of COVID-19, and lockdown to prevent the spread of COVID-19. Regarding the assessment of practice, the question was composed of 3 questions (P1-P3): the idea of grocery stocking, preventive measures during the lockdown, and relationship with family and friends. Similar scoring pattern as knowledge was kept. Cronbach’s alpha was used to measure the internal consistency of the questionnaire.

### Statistical analysis

The data were analysed via statistical Package for the Social Sciences (IBM Corp. Released 2013. IBM SPSS Statistics for Windows, Version 22 Armonk, Chicago, Illinois: IBM Corp). Mean with standard deviation was calculated for descriptive analysis and the number with percentage was calculated for categorical variables. Knowledge, attitude, and practice score were compared by demographics with chi-square or Fischer’s exact test as appropriate. Multivariate linear regression analysis was used to establish the relationship between demographic variables as independent variables and KAP as the outcome variable. Pearson Coefficient of correlations and VIF (Variance of inflation factor) were used to determine the relationship between knowledge, attitude, and practice. Unstandardized regression coefficient (β) with a 95% Confidence interval was used to quantify the relation between variables and KAP. Statistical Significance was set at ≤ 0.05.

#### Ethical Clearance

The study was cleared by the Institutional Ethical Committee, Nims Medical College, Nims University Rajasthan, Jaipur.

## RESULTS

### Study population characteristics

A total of 8075 questionnaires were retrieved, of which 97 questionnaires were excluded as the participants declined to give consent. The remaining 7978 questionnaires were completed with an overall response rate of 98.79%. The details of demographic characteristics were presented in (Table 2). A large proportion of male respondents (52.4%) were found. The leading age band was 18-30 years, accounting approximately 54.2% of both the genders. Additionally, Total of 5588 (70.04%) subjects holds a degree of graduate or above, 3232 (40.5%) pursues as a student or unemployed and 5108 (64%) were single.

**Table-1.** KAP of COVID-19 among7978 respondents, India n (%)

| S.NO | QUESTIONS |  |  |  |
| --- | --- | --- | --- | --- |
|  | KNOWLEDGE | YES | NO | NOT CLEAR |
| K1. | COVID-19 spread through: contact to an infected person, respiratory droplets, and oral-fecal route. (91.97%)* | 7338(91.7%) | 494(6.19%) | 147(1.84%) |
| K2. | The early sign of COVID-19 is fever, difficulty in breathing, cough, and sore throat. (95.97%)* | 7657(95.98%) | 223(2.79%) | 98(1.23%) |
| K3. | Do you agree washing hands on a regularly is an effective measure to the kill the novel Coronavirus? (97.90%) | 7814(97.92) | 157(1.96%) | 7(0.10) |
| K4. | Do you think that risk of spread of novel Coronavirus is from both symptomatic as well as non-symptomatic cases? (91.61%)* | 7309(91.61%) | 514(6.44%) | 156(1.95%) |
| K5. | Symptomatic treatment, early screening, and social distancing are important measure to stop the spread of the novel Coronavirus. (97.05%)* | 7743(97.05%) | 39(0.49%) | 196(2.46%) |
| K6. | The disease is more dangerous in people with cancer, diabetes, and chronic respiratory disease? (86.11%)* | 6871(86.12%) | 513(6.43%) | 594(7.45%) |
| K7. | Do you agree that isolation and quarantine are an important step to stop the spread of novel Coronavirus? (89.4%)* | 7134(89.42%) | 529(6.63%) | 315(3.95%) |
| K8. | An Isolation period of 14 days is important for a person who had contact with Coronavirus -infected patients? (95.15%)* | 7592(95.16%) | 245(3.07%) | 141 (1.77%) |
| K9. | Currently, there is no specific vaccine and cure for COVID-19. However, isolation, early symptomatic and supportive treatment can help patients recover from the infection? (88.99%)* | 7101(88.99%) | 304(3.81) | 573(7.20%) |
| K10. | Do you think pregnant women are more at risk towards COVID-19? (83.14%) * | 6633(83.14%) | 349(4.37%) | 996(12.49%) |
| K11. | Do you think that eating non-vegetarian food is not responsible for spread the of COVID-19? (96.3%)* | 7687(96.35%) | 95(1.19%) | 196(2.46%) |
| K12. | Do you agree that eating citrus fruit and gargling with salt water can help prevent infection with the novel Coronavirus? (37%)* | 5024(62.97) | 2818(35.31%) | 136(1.70%) |
| K13. | Do you think that drinking alcohol can prevent you from COVID-19? (86%)* | 1118(14.02%) | 5152(64.58%) | 1708(21.40) |
|  | ATTITUDE | YES | NO |  |
| A1. | In recent days, have you gone to any populated place? (97.63%)* | 189(2.36%) | 7789(97.63%) |  |
| A2. | Are you confident that the world will win the battle against COVID-19? (98.34%)* | 7846(98.35%) | 132(1.65%) |  |
| A3. | Do you agree with the idea of lockdown to prevent the spread of COVID -19? (96.03%)* | 7661(96.03%) | 317(3.97%) |  |
|  | PRACTICE | YES | NO |  |
| P1. | Do you agree with the idea of grocery and medicine stocking? (59.42)* | 3236(40.56%) | 4742(59.43%) |  |
| P2. | Do you take proper preventive measures while leaving home? (94.04)* | 7503(94.05%) | 475(5.95%) |  |
| P3. | Do you feel more bonded to your family and friends in the duration of lockdown? (97.98%)* | 7818(97.99%) | 160(2.01%) |  |

\*Percentage of correct answer

**Table-2.** Characteristics of study of participants (n=7978)

| CHARACTERISTICS | *n=7978(%) |
| --- | --- |
| **GENDER**<br>Female<br>Male | 3795 (47.6)<br>4183 (52.4) |
| **AGE**<br>18-30 Years<br>30-50 Years<br>>50 Years | 4328 ( 54.2)<br>2354 (29.5)<br>1296 (16.2) |
| **MARITAL STATUS**<br>Married<br>Single<br>Others | 2656 (33.3)<br>5108 (64)<br>214 (2.7) |
| **GEOGRAPHICAL AREA**<br>Rural<br>Urban | 3568 (44.7)<br>4410 (55.3) |
| **EDUCATION**<br>Below secondary school<br>Senior secondary<br>Graduate<br>Post-graduate and above | 350 (4.4)<br>2039 (25.6)<br>2802 (35)<br>2787 (34.9) |
| **OCCUPATION**<br>Unskilled<br>Skilled<br>Student and Unemployed<br>Self employed<br>Professional | 298 (3.7)<br>313 (3.9)<br>3232 (40.5)<br>2027 (25.4)<br>2108 (26.4) |

\*n= Total number of population

### Knowledge Score related to COVID-19

As to knowledge, the mean was 11.36 ± 1.2 (range 0-13) suggesting an overall 80.64% correct rate of knowledge. Univariate analysis with knowledge level significantly varies across age, gender, education, and occupation. 89.4% agreed that isolation and quarantine are an important step to stop the spread of novel coronavirus and only 83.19% of respondents were aware of pregnant women are more at risk towards COVID-19. Whereas, only 37% believed eating citrus fruit and gargling with salt water cannot help in preventing infection with the Nobel Coronavirus (Table 3). In multiple linear regression analysis, male (β=0.036: p<0.001), urban population (β=0.006:p<0.002), higher education (β=0.029:p<0.001), higher occupation (β=0.002:p=0.05) have associated significantly with high knowledge score. There is no evidence of multicollinearity between independent variables (range of VIP 1.102 and 1.287) (Table 4).

**Table-3.** Correlation of demographic and the knowledge based on univariate analysis.

| KNOWLEDGE* |  |  |  |  |  |  |  |
| --- | --- | --- | --- | --- | --- | --- | --- |
| Socio-demographic characteristics | Total Score (mean±SD) | Level(ROW%), n=7978 |  | X<sup>2</sup>-value | P-value | F(exact) | df*** |
|  |  | Excellent** | Poor |  |  |  |  |
| **GENDER**<br>Female<br>Male | 11.04 ± 1.05<br>11.65 ± 1.29 | 73.5%<br>87.1% | 26.5%<br>12.9% | 234.590 | <0.001 | .000 | 1 |
| **AGE**<br>18-30 Years<br>30-50 Years<br>>50 Years | 11.17 ± 1.30<br>11.53 ± 1.07<br>11.69 ± 1.03 | 74%<br>86%<br>93.1% | 26%<br>14%<br>6.9% | 294.400 | <0.001 | - | 2 |
| **MARITAL STATUS**<br>Married<br>Single<br>Other<sup>#</sup> | 12.47 ±0.86<br>11.11 ± 1.24<br>10.59 ± 1.05 | 73.3%<br>85.6%<br>53.7% | 26.7%<br>14.4%<br>46.3% | 0.060 | 0.971 | - | 2 |
| **GEOGRAPHICAL AREA**<br>Rural<br>Urban | 11.30 ± 1.22<br>11.42 ±1.21 | 79.6%<br>81.4% | 20.4%<br>18.6% | 4.360 | 0.390 | 0.04 | 1 |
| **EDUCATION**<br>Below secondary school<br>Senior secondary<br>Graduate<br>Post-graduate and above | 10.00 ± 1.48<br>11.81 ± 0.87<br>11.63 ± 1.51<br>11.63 ± 0.94 | 41.7%<br>56.9%<br>90.4%<br>93% | 58.3%<br>43.1%<br>9.6%<br>7% | 1518.600 | <0.001 | - | 3 |
| **OCCUPATION**<br>Unskilled<br>Skilled<br>Student and Unemployed<br>Self employed<br>Professional | 10.29 ± 1.46<br>11.14 ± 1.40<br>11.40 ± 1.12<br>11.11 ± 1.37<br>11.43 ± 1.21 | 45.6%<br>72.2%<br>81.8%<br>70.0%<br>95.1% | 54.4%<br>27.8%<br>18.2%<br>30%<br>4.9% | 679.980 | <0.001 | - | 4 |

\*The questions and options of each questions are describe in Table 1.
\*\*Knowledge score cut-off of ≥80% is used to define excellent knowledge.
\*\*\*df- degree of freedom
<sup>#</sup> "Others" include re-married, separated, divorced, and widowed.
P<0.05 statistically significant

**Table-4.** Participants characteristics with knowledge score regarding COVID-19 based on multiple linear regression.

| **KNOWLEDGE\*** |  |  |  |  |  |
| --- | --- | --- | --- | --- | --- |
| **Socio-demographic characteristics** | **Unstandardised coefficients(B)** | **Standardised coefficients(B)** | **P-value** | **95% coefficient interval(B)** | **VIF\*\*** |
| **GENDER**<br>Female<br>Male | 0.036 | 0.194 | <0.001 | 0.032 to 0.040 | 1.109 |
| **AGE**<br>18-30 Years<br>30-50 Years<br>>50 Years | 0.001 | 0.007 | 0.550 | -0.002 to 0.004 | 1.287 |
| **MARITAL STATUS**<br>Married<br>Single<br>Others<sup>#</sup> | 0.004 | 0.020 | 0.060 | 0.000 to 0.008 | 1.102 |
| **GEOGRAPHICAL AREA**<br>Rural<br>Urban | 0.006 | 0.033 | 0.002 | 0.002 to 0.010 | 1.052 |
| **EDUCATION**<br>Below secondary school<br>Senior secondary<br>Graduate<br>Post-graduate and above | 0.029 | 0.275 | <0.001 | 0.027 to 0.032 | 1.516 |
| **OCCUPATION**<br>Unskilled<br>Skilled<br>Student and Unemployed<br>Self employed<br>Professional | 0.002 | 0.023 | 0.050 | 0.000 to 0.004 | 1.310 |

*\*The questions and options of each questions are described in Table 1. \*\*VIF- Variance inflation factor # “Others” include re-married, separated, divorced, and widowed. P<0.05 statistically significant*
*\*The questions and options of each questions are described in Table 1. \*\*VIF- Variance inflation factor

### Attitude Score related to COVID-19

The overall correct rate in attitude was 97.33%. Majority of population 97.6% had not been any populated place. Nearly respondents (98.33%) believed that COVID-19 can be successfully controlled. Moreover, 96.01% agreed with the idea of lockdown to prevent the spread of COVID-19. Univariate analysis is significantly associated with age, gender, marital status, area of residence, education, and occupation (Table 5). In multiple linear regression analysis, female(β=-0.006:p=0.008), middle-age person (β=-0.011:p<0.001), higher education (β=0.025:p<0.001), higher occupation (β=0.020:p<0.001) have associated significantly with good attitude score. There is no evidence of multicollinearity between marital status, geographical area, and attitude (range of VIP = 1.102 and 1.052 p>0.05) (Table 6)

**Table-5.** Correlation of demographic and the attitude based on univariate analysis.

| **ATTITUDE\*** |  |  |  |  |  |  |  |  |  |
| --- | --- | --- | --- | --- | --- | --- | --- | --- | --- |
| **Socio-demographic characteristics** | **A1. In recent days, have you gone to any populated place?** |  |  | **A2. Are you confident that world will win the battle against COVID-19?** |  |  | **A3. Do you agree with the idea of lockdown to prevent the spread of COVID-19?** |  |  |
|  | **YES**<br>**(n, Row %)** | **NO**<br>**(n, Row %)** | **X<sup>2</sup>-value**<br>**p-value**<br>**df\*\*** | **YES**<br>**(n, Row %)** | **NO**<br>**(n, Row %)** | **X<sup>2</sup>-value**<br>**p-value**<br>**df\*\*** | **YES**<br>**(n, Row %)** | **NO**<br>**(n, Row%)** | **X<sup>2</sup>-value**<br>**p-value**<br>**df\*\*** |
| **GENDER**<br>Female (47.6%)<br>Male (52.4%) | 40(1.1%)<br>149(3.6%) | 3755(98.9%)<br>4034(96.4%) | X<sup>2</sup>=54.130<br>p<0.001<br>df=1 | 3740(98.6%)<br>4106(98.2%) | 55(1.4%)<br>77(1.8%) | X<sup>2</sup>=1.875<br>p=1.880<br>df=1 | 3580(94.3%)<br>4081(97.6%) | 215(5.7%)<br>102(2.4%) | X<sup>2</sup>=54.240<br>p<0.001<br>df=1 |
| **AGE**<br>18-30 Years (54.2%)<br>30-50 Years (29.5%)<br>>50 Years (16.2%) | 120(2.8%)<br>33(1.4%)<br>36(2.8%) | 4208(97.2%)<br>2321(98.6%)<br>1260(97.2%) | X<sup>2</sup>=13.500<br>p=0.001<br>df=2 | 4237(97.9%)<br>2326(98.8%)<br>1283(99%) | 91(2.1%)<br>28(1.2%)<br>13(1%) | X<sup>2</sup>=11.840<br>p=0.003<br>df=2 | 4109(94.9%)<br>2300(97.7%)<br>1252(96.6%) | 219(5.1%)<br>54(2.3%)<br>44(3.4%) | X<sup>2</sup>=31.930<br>p<0.001<br>df=2 |
| **MARITAL STATUS**<br>Married (33.3%)<br>Single (64%)<br>Others<sup>#</sup> (2.7%) | 56(2.1%)<br>109(2.1%)<br>24(11.2%) | 2600(97.9%)<br>4999(97.9%)<br>190(88.8%) | X<sup>2</sup>=74.400<br>p<0.001<br>df=2 | 2608(98.2%)<br>5035(98.6%)<br>203(94.9%) | 48(1.8%)<br>73(1.4%)<br>11(5.1%) | X<sup>2</sup>=17.950<br>p<0.001<br>df=2 | 2490(93.8%)<br>4970(97.3%)<br>201(93.9%) | 166(6.3%)<br>138(27%)<br>13(6.1%) | X<sup>2</sup>=60.200<br>p<0.001<br>df=2 |
| **GEOGRAPHICAL AREA**<br>Rural (44.7%)<br>Urban (55.3%) | 80(2.2%)<br>109(2.5%) | 3488(97.8%)<br>4301(97.5%) | X<sup>2</sup>=0.449<br>p=0.500<br>df=1 | 3505(98.2%)<br>4341(98.4%) | 63(1.8%)<br>69(1.6%) | X<sup>2</sup>=0.490<br>p=0.484<br>df=1 | 3400(95.3%)<br>4261(96.6%) | 168(4.7%)<br>149(3.4%) | X<sup>2</sup>=9.140<br>p=.002<br>df=1 |
| **EDUCATION**<br>Below secondary school (4.4%)<br>Senior secondary (25.6%)<br>Graduate (35.1%)<br>Post-graduate and above (34.9%) | 31(8.9%)<br>69(3.4%)<br>73(2.6%)<br>16(0.6%) | 319(91.1%)<br>1970(96.6%)<br>2729(97.4%)<br>2771(99.4%) | $X^2=112.280$<br>$p<0.001$<br>df=3 | 346(98.9%)<br>2013(98.7%)<br>2722(97.1%)<br>2765(99.2%) | 4(1.1%)<br>26(1.3%)<br>80(2.9%)<br>22(0.8%) | $X^2=40$<br>$p<0.001$<br>df=3 | 162(46.3%)<br>1935(94.9%)<br>2792(99.6%)<br>2772(99.5%) | 188(53.7%)<br>104(5.1%)<br>10(0.4%)<br>15(0.5%) | $X^2=2458$<br>$p=0.001$<br>df=3 |
| **OCCUPATION**<br>Unskilled (3.7%)<br>Skilled (3.9%)<br>Student and Unemployed (40.5%)<br>Self employed (25.4%)<br>Professional (26.4%) | 23(7.7%)<br>21(6.7%)<br>43(1.3%)<br>51(2.5%)<br>51(2.4%) | 275(92.3%)<br>292(93.3%)<br>3189(98.7%)<br>1976(97.5%)<br>2057(97.6%) | $X^2=77.640$<br>$p=0.001$<br>df=4 | 295(99.0%)<br>307(98.1%)<br>3147(97.4%)<br>2022(99.8%)<br>2075(98.4%) | 3(1.0%)<br>6(1.9%)<br>85(2.6%)<br>5(0.2%)<br>33(1.6%) | $X^2=44.590$<br>$p<0.001$<br>df=4 | 117(39.3%)<br>211(67.4%)<br>3217(99.5%)<br>2016(99.5%)<br>2100(99.6%) | 181(60.7%)<br>102(32.6%)<br>15(0.5%)<br>11(0.5%)<br>8(0.4%) | $X^2=3426$<br>$p<0.001$<br>df=4 |

*\*The questions and options of each question are described in Table 1.* *\*\*df- degree of freedom* *# "Others" include re-married, separated, divorced, and widowed.* *P<0.05 statistically significant*

**Table-6.**
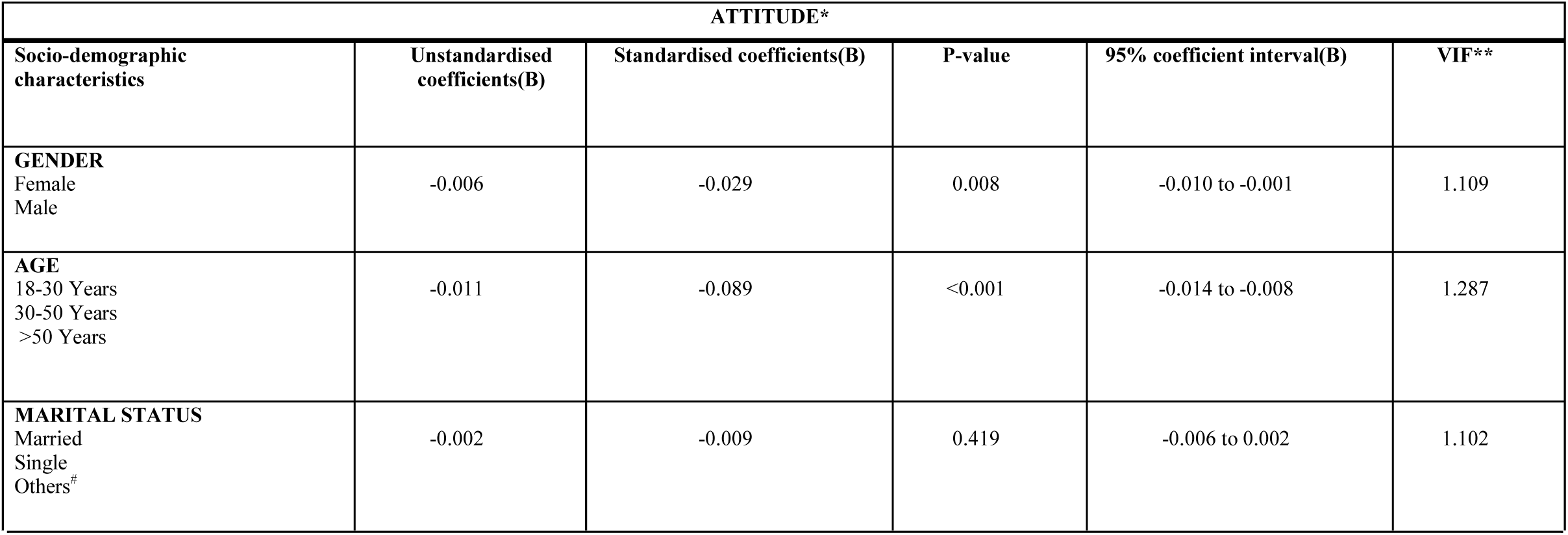
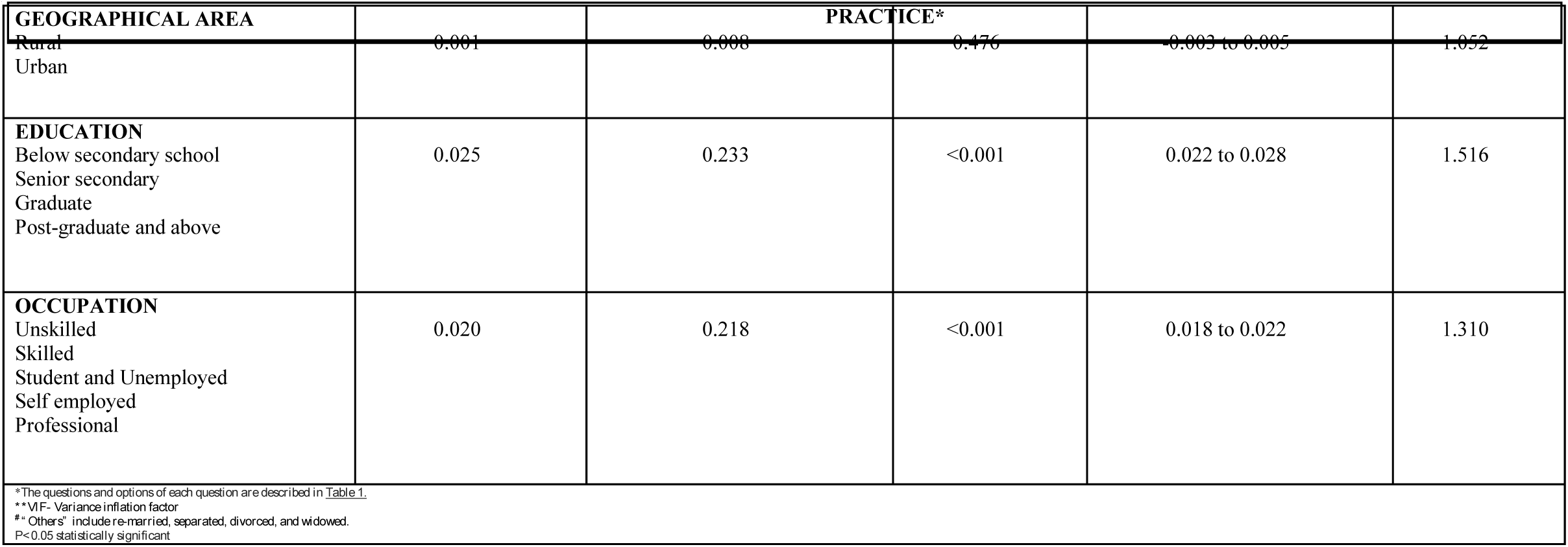
Participants characteristics with attitude score regarding COVID-19 based on multiple linear regression.

### Practice Score related to COVID-19

Based on results overall response rate was 83.8 %. In the study, most males (72.7%) denied the idea of grocery stocking in contrast, 55.2% of females agree with this idea of grocery stocking. Both the gender (90.7%) males and (97.1%) taking proper preventive measures while leaving home (Table 7) In multiple regression, male (β=0.093:p<0.001), old age (β=0.030:p<0.001), single and other (β=0.113:p<0.001), lower education (β=-0.007:p=0.007) have associated significantly with good practice. However, there is no evidence of multicollinearity of geographical area and occupation with practice score (range of VIP =1.052 and 1.310 respectively p<0.05) (Table 8).

**Table-7.** Correlation of demographic and the practice based on univariate analysis.

| Socio-demographic characteristics | P1. Do you agree with the idea of grocery and medicine stocking? |  |  | P2.Do you take proper preventive measures while leaving home? |  |  | P3.Do you feel more bounded to your family and friends in the duration of lockdown? |  |  |
| --- | --- | --- | --- | --- | --- | --- | --- | --- | --- |
|  | YES<br>(n, Row %) | NO<br>(n, Row %) | X<sup>2</sup>-value<br>p-value<br>df** | YES<br>(n, Row%) | NO<br>(n, Row%) | X<sup>2</sup>-value<br>p-value<br>df** | YES<br>n, (Row %) | NO<br>(n, Row %) | X<sup>2</sup>-value<br>p-value<br>df** |
| **GENDER**<br>Female (47.6%)<br>Male (52.4%) | 2095(55.2%)<br>1141(27.3%) | 1700(44.8%)<br>3042(72.7%) | X<sup>2</sup>=643<br>p<0.001<br>df=1 | 3443(90.7%)<br>4059(97.1%) | 352(9.3%)<br>123(2.9%) | X<sup>2</sup>=142.500<br>p<0.001<br>df=1 | 3666(96.6%)<br>4151(99.3%) | 129(3.4%)<br>31(0.7%) | X<sup>2</sup> = 71.510<br>p<0.01<br>df=1 |
| **AGE**<br>18-30 Years (54.2%)<br>30-50 Years (29.5%)<br>>50 Years (16.2%) | 1976(45.7%)<br>828(35.2%)<br>432(33.3%) | 2352(54.3%)<br>1526(64.8%)<br>864(66.7%) | X<sup>2</sup>=103<br>p<0.001<br>df=2 | 3976(91.9%)<br>2271(96.5%)<br>1218.8(96%) | 352(8.1%)<br>83(3.5%)<br>40(3.1%) | X<sup>2</sup> = 80.540<br>p<0.001<br>df=2 | 4212(97.3%)<br>2320(98.6%)<br>1286(99.2%) | 116(2.7%)<br>34(1.4%)<br>10(0.8%) | X<sup>2</sup>=23.830<br>p<0.001<br>df=2 |
| **MARITAL STATUS**<br>Married (33.3%)<br>Single (64%)<br>Others<sup>#</sup> (2.7%) | 1846(69.5%)<br>1326(26%)<br>64(29.4%) | 810(30.5%)<br>3782(74%)<br>150(70.1%) | X<sup>2</sup>= 1384<br>p=<0.001<br>df=2 | 2504(94.3%)<br>4786(93.7%)<br>213(99.5%) | 152(5.7%)<br>322(6.3%)<br>1(0.2%) | X<sup>2</sup> = 12.870<br>p=0.002<br>df=2 | 2593(97.6%)<br>5011(98.1%)<br>214(100%) | 63(2.4%)<br>97(1.9%)<br>0 | X<sup>2</sup> = 6.490<br>p=0.039<br>df=2 |
| **GEOGRAPHICAL AREA**<br>Rural (44.7%)<br>Urban (55.3%) | 1596(44.7%)<br>1640(37.2%) | 1972(55.3%)<br>2770(34.7%) | X<sup>2</sup> = 46.540<br>p=<0.001<br>df=1 | 3347(93.8%)<br>4156(94.2%) | 221(6.2%)<br>254(5.8%) | X<sup>2</sup> = 0.664<br>p=0.410<br>df=1 | 3501(98.1%)<br>4317(97.9%) | 97(1.9%)<br>93(2.1%) | X<sup>2</sup>= 0.536<br>p=0.46<br>df=1 |
| **EDUCATION**<br>Below secondary school (4.4%)<br>Senior secondary (25.6%)<br>Graduate (35.1%)<br>Post-graduate and above (34.9%) | 178(50.9%)<br>852(41.8%)<br>1229(43.9%)<br>977(35.1%) | 172(49.1%)<br>1187(58.2%)<br>1573(56.1%)<br>1810(64.9%) | X<sup>2</sup>=64.350<br>p-<0.001<br>df=3 | 333(95.1%)<br>1935(94.9%)<br>2557(91.3%)<br>2678(96.1%) | 17(4.9%)<br>104(5.1%)<br>245(8.7%)<br>109(3.9%) | X<sup>2</sup>= 63.01<br>p<0.001<br>df=3 | 345(98.6%)<br>2003(98.2%)<br>2705(96.5%)<br>2765(99.2%) | 5(1.4%)<br>36(1.8%)<br>97(3.5%)<br>22(0.8%) | X<sup>2</sup> = 52.40<br>p<0.01<br>df=3 |
| **OCCUPATION**<br>Unskilled (3.7%)<br>Skilled (3.9%)<br>Student and Unemployed (40.5%)<br>Self employed (25.4%)<br>Professional (26.4%) | 154(51.7%)<br>121(38.7%)<br>1282(39.7%)<br>851(42%)<br>828(39.3%) | 144(48.3%)<br>192(61.3%)<br>1950(60.3%)<br>1176(58%)<br>1280(60.7%) | X<sup>2</sup>- = 19.950<br>p=0.001<br>df=4 | 285(95.6%)<br>299(95.5%)<br>2997(92.7%)<br>1888(93.1%)<br>2034(96.5%) | 13(4.4%)<br>14(4.5%)<br>235(7.3%)<br>139(6.9%)<br>74(3.5%) | X<sup>2</sup> = 38.020<br>p<0.001<br>df=4 | 298(100%)<br>307(98.1%)<br>3142(97.2%)<br>1977(97.5%)<br>2094(99.3%) | 0(0%)<br>6(1.9%)<br>90(2.8%)<br>50(2.5%)<br>14(0.7%) | X<sup>2</sup>= 37.580<br>p<0.01<br>df=4 |

\*The questions and options of each questions are described in Table 1.
\*\*df- degree of freedom

**Table-8.** Participants characteristics with practice score regarding COVID-19 based on multiple linear regression.

| PRACTICE* |  |  |  |  |  |
| --- | --- | --- | --- | --- | --- |
| Socio-demographic characteristics | Unstandardised coefficients(B) | Standardised coefficients(B) | P-value | 95% coefficient interval(B) | VIF** |
| **GENDER**<br>Female<br>Male | 0.093 | 0.234 | <0.001 | 0.084 to 0.101 | 1.109 |
| **AGE**<br>18-30 Years<br>30-50 Years<br>>50 Years | 0.030 | .114 | <0.001 | 0.024 to 0.036 | 1.287 |
| **MARITAL STATUS**<br>Married<br>Single<br>Others<sup>#</sup> | 0.113 | 0.295 | <0.001 | -0.006 to 0.010 | 1.102 |
| **GEOGRAPHICAL AREA**<br>Rural<br>Urban | 0.002 | 0.004 | 0.691 | -0.006 to 0.010 | 1.052 |
| **EDUCATION**<br>Below secondary school<br>Senior secondary (25.6%)<br>Graduate (35.1%)<br>Post-graduate and above (34.9%) | -0.007 | -0.033 | 0.007 | -0.013 to -0.002 | 1.516 |
| **OCCUPATION**<br>Unskilled (3.7%)<br>Skilled (3.9%)<br>Student and Unemployed<br>Self employed<br>Professional | -0.002 | -0.009 | 0.441 | -0.006 to 0.003 | 1.310 |

\*The questions and options of each questions are described in Table 1.
\*\*VIF- Variance inflation factor
<sup>#</sup> "Others" include re-married, separated, divorced, and widowed.
P<0.05 statistically significant

### Correlation between knowledge, attitude, and practice scales

There was positive significant correlation between knowledge-attitude, knowledge-practice and attitude practice with strongest relation identified between practice and attitude. Express by Pearson’s correlation, the knowledge score correlation with Attitude (r=0.023:p<0.01) and practice (r=0.019:p<0.01). Moreover, practice score positive correlation with attitude (r=0.361:p=0.03). (Table 9)

**Table-9.** Correlation between score of knowledge, attitude, practice.

|  | CORRELATION | P-VALUE |
| --- | --- | --- |
| Knowledge- Attitude | 0.153* | <0.001 |
| Knowledge- Practice | 0.139* | <0.001 |
| Attitude- Practice | 0.101* | 0.037 |
| *\*Correlation is significant at the 0.01 level(2-tailed)*<br>*P<0.05 statistically significant* |  |  |

## DISCUSSION

Since the outbreak, Corona virus had brought chaos to lives and economics around the world. As of now, India is facing biggest health emergency since the country has gained independence. Encouragement of public to adopt precautionary behaviours for containment strategies as knowledge attitude and practice is foreground for public cooperation and backbone for implementing any health policy. Countries have reported community response to COVID-19 from china and middle east [20,24]. From the aspect of attention, this is the unique paper from India including 7978 subjects analysed towards COVID-19.

Based on our findings, the study significantly consists males, single and well-educated population. The overall 80.64% of knowledge score was higher and unanticipated. Essentially because the online survey was after the ubiquitous mass media coverage on rampant caused by COVID-19 to global giants i.e (America, Europe, China) and through effective health communication for sustainable adoption of preventive measures by Ministry of Health & family welfare, Government of India [25,26]. Furthermore, Sample Characteristics: consisting participants with higher education holds the predication of knowledge Score which is similar to Chinese study but varied with Iranian study [20,24], Participants were aware regarding clinical symptoms, transmission, prevention and control of disease contrast to myth busters questions. More than one-third participants agreed about eating citrus fruits and gargling salt water can help prevent infection. According to WHO gargling warm or saltwater and consuming citrus fruits will not kill novel-corona virus [27]. 14% out of hundred think alcohol drinking can kill novel coronavirus. Alcohol are highly inflammable substance as well as strong disinfectants, it can be used as a cleansing agent for surfaces [28]. No evidence supports alcohol consumption can kill the virus, whereas intake of excessive alcohol can cause health related complications. Robust association between gender and knowledge score regarding COVID-19 was found in the present study. Multivariate analysis showed confounding factors (education level and occupation) as strong indicator of knowledge domain regarding COVID-19 which postulates the combination of better access to information and high education level leads to appropriate apprehension and comprehension of information on covid-19, consequent to better knowledge on COVID-19. Therefore, government and public health policy-makers should recognize target populations for COVID-19 prevention and health education.

Our study has strong association of knowledge significantly with positive attitude and practice. In recent, majority of population (97.6%) have taken optimistic measures by avoiding going to crowded places. This practice is primarily due to strict measures taken by government to prevent overwhelmed wave of new infection whereas secondly due to awareness, acceptance and action of people with good knowledge. Unfortunately, 3.6% males are going to crowded place because of two potential reason: first in India men are likely to expose in outwork exposing them to crowded place, secondly according to studies men and adolescents have more risk-taking behaviour which suggestive of dangerous practice towards COVID-19. Similar findings were obtained in previous studies conducted in china [20]. Massive agreement was found towards the idea of lockdown in India, which has helped to prevent potential rise in cases. According to ICMR (Indian medical Council of Research): Reproduction factor (Ro) of SARS-CoV-2 is 2.5. One person can infect 406 peoples in 30 days, if the lockdown and social distancing are placed properly one sick person can only infect 2.5 persons. Successful containment of pandemic is social distancing which is already proven by millennial example in pandemics of influenza [29,30,31].

Lack of information, imprecise information, deception can lead to hysteric and fuel behavioural outcomes for example – panic buying [32]. According to recent studies, Panic buying is having detrimental impact on health supply chains leading to shortage of essentials like sanitizers, masks and pain relievers [33]. In our study, 55% of females and 27.3 % males agreed with idea of grocery and medicine stocking. Multiple Linear regression suggested married and age significantly associated in stocking practice. As panic buying disturbs the balance of demand and supply in goods supply chain system [34], no research was found to establish relation between association of marital status and gender in grocery and medicine stocking. Mental health is another major issue that is becoming critical in managing COVID-19 pandemic. Social chaos and arbitrary relationship are destroyed due to panic and fear, thus superseding evidence and Psychologically, change in environment makes us feel unsafe, scared and anxious. Family and friends are major source to mental ability by reassurance well-being and high spirits [35, 36]. In our study predominant population experience connected with family and friends [37].

Strength of the study is large sample size during the critical period of nationwide lockdown and COVID-19 outbreak. Our sample was perfectly balanced as both the genders were recruited which really made us to take effort as in India more telecom users are males Considering that educational attainment and occupation are frequently considered proxy measures od socio-economic status. Significant association between demographics variables and KAP towards COVID-19, due to which we have overestimated rates of preventive practice and attitudes with the actual practice.

Due to limited access of internet and proper health related resources rural people and aged adults with unskilled occupation have poor knowledge. Another limitation to the study is limited question in attitude and practice due to limited time for developing questionnaire.

## CONCLUSION

To conclude, majority of Indian population demonstrated preceded good knowledge, positive attitude and good practice regarding COVID-19 pandemic. Furthermore, due to systemic approach and health communication strategies significant awareness and apprehension in knowledge, preventive strategies and optimistic attitude was assimilated general population. Other than that, government policy makers has worked well in targeting grass-root population having low level of education and non-professional workers and managed proactively the concept of “social distancing” in COVID-19.

Noteworthily India in unison with strengthen health systems measures and cooperation with public-health policy makers will fight back this pandemic with optimistic control and empowered knowledge.

**Figure-1.**
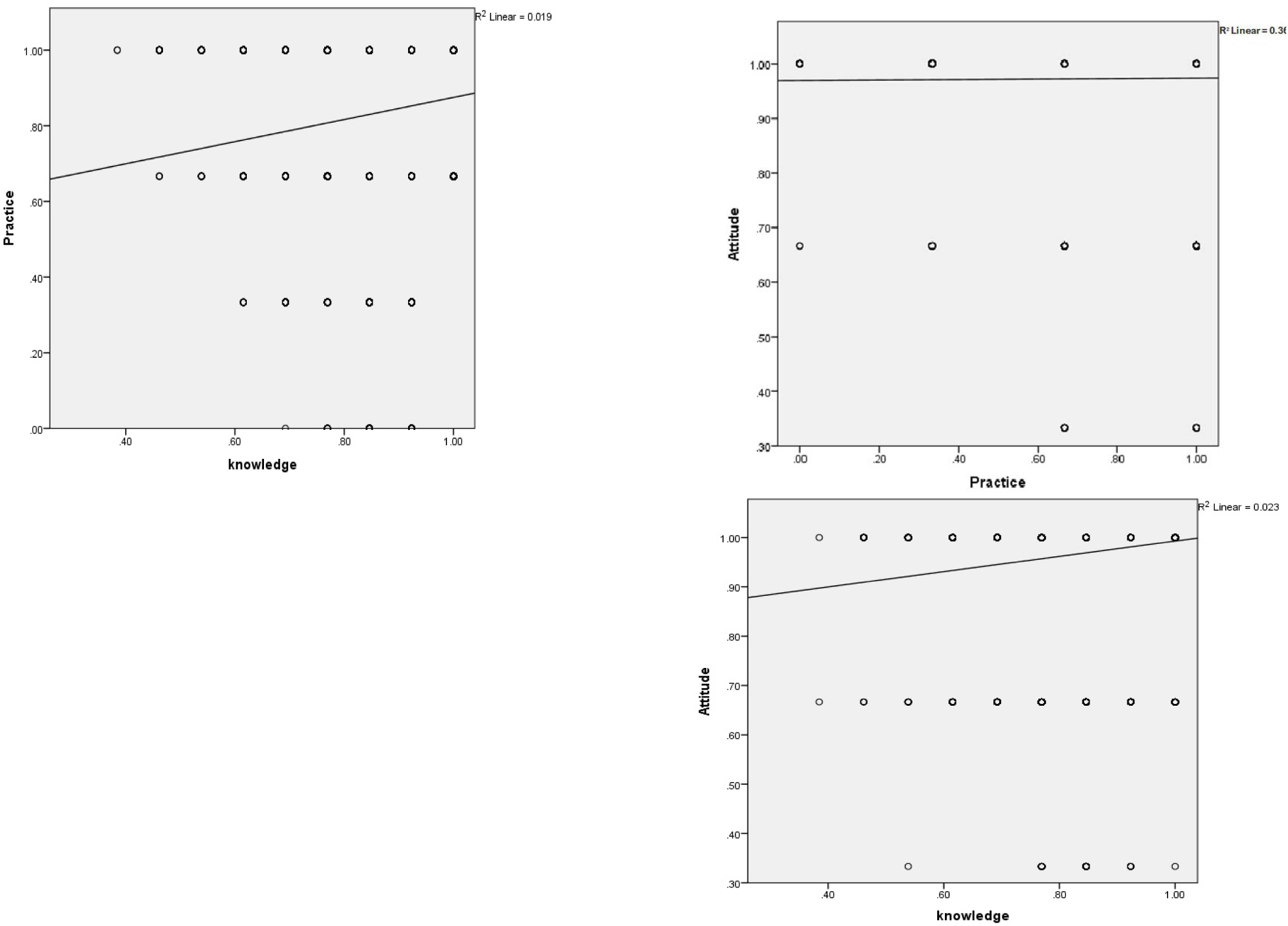
Correlation scattered among knowledge, attitude, and practice in Indian general population.

## Data Availability

All the data provided are derived from original research and true in nature.

